# Psilocybin Experiential Therapist Training: Insights from a World-First Study

**DOI:** 10.1101/2025.11.15.25340324

**Authors:** Georgia Ioakimidis-MacDougall, John Gardner, Paul Liknaitzky

## Abstract

First-hand experience with psychedelics may help clinicians develop skills and knowledge needed to work with the profound changes to conscious awareness occasioned by psychedelics. However, the topic remains contentious and underexplored. In this world-first study, we investigated the utility of psilocybin experiential therapist training in a sample of 14 mental healthcare professionals training to provide psilocybin-assisted therapy. Participants received one 25 mg dose of psilocybin in a clinical research context alongside psychological support before, during, and after dosing. Quantitative measures and semi-structured interviews were then undertaken by participants to explore their experiences and reflections. Through the intervention, participants reported developing a greater and embodied understanding of key therapeutic principles and processes. Moreover, they reported increases in therapeutic qualities (e.g., empathy, attunement, emotion regulation) that underpin therapeutic alliance and promote trust and safety. While participants did not report experiencing harms from participation, they speculated about two potential risks of psychedelic experiential therapist training: first, that it could elicit challenging material that feels destabilising for a period; and second, that therapists could project their experience onto clients in a manner that narrows interpretative range and reduces attunement. Recommendations were made for psychedelic experiential therapist training design and implementation, including strategies to mitigate such risks. Participants indicated that psychedelic experiential therapist training is necessary but not sufficient for providing the highest quality of care in psychedelic-assisted therapy. Findings support the inclusion of an optional psychedelic experiential component within psychedelic therapist training programs for clinicians with prior psychotherapeutic training and well-developed reflective capacity.

## Introduction

Growing empirical evidence highlights the potential of psilocybin-assisted therapy (PAT) for treating a range of mental health conditions, including unipolar depression, substance use disorders, and end-of-life distress (Carhart-Harris et al., 2021; Griffiths et al., 2016; Johnson et al., 2017; Raison et al., 2023). Classic psychedelics, including psilocybin, produce profound changes to waking consciousness that can be entirely unfamiliar, ontologically shocking, and ineffable (i.e., beyond the descriptive capacity of language; Sanders & Zijlmans, 2021). Given this, many researchers and clinicians espouse the importance of psychedelic therapists having first-hand experience with psychedelics to develop the empathy, knowledge, and skills necessary to work therapeutically with them (Dames et al., 2024; Halberstadt, 2014; McLaughlin & Grof, 1976; Passie et al., 2025). However, in contemporary psychiatry, prescribers are not expected to have personal experience with the psychotropic medications they prescribe. Further, the use of illicit substances such as psychedelics by researchers and mental healthcare professionals remains stigmatised (Nielson & Guss, 2018). Due in part to these factors, questions around the utility of psychedelic experiential learning are under-explored, and psychedelic experiential therapist training remains largely excluded from above-ground psychedelic therapist training programs (Dames et al., 2024). The resurgence of clinical trials of PAT globally, the early clinical access in some jurisdictions like Australia, and the near-term prospect of approved psychedelic medicines has made the need for training standards, competency frameworks, and larger-scale training programs clear (Nutt & Carhart-Harris, 2021). Relevant to the key question of how best to train psychedelic therapists, this study aimed to explore the potential utility of using psilocybin with support as a training tool.

Direct experience with psychedelics has historically been valued by those who work therapeutically with these substances. For thousands of years, indigenous healing practices around the world involving plant-based psychedelics have typically been led by practitioners with extensive personal psychedelic experience (George et al., 2019). During an initial period of clinical psychedelic research (1950s-1970s), it was common for psychedelic researchers and clinicians to have personal experience with the substances they worked with (Sessa, 2012). Knowledge gained from such experiences was thought to assist with understanding, interpreting, and supporting psychedelic phenomena in their patients (Hoffman, 1980). Conversely, some early psychedelic therapists suggested that without such exposure, clinicians were less equipped to support patients during sessions or interpret the subtleties of their experience (Mangini, 1998). In contemporary discourse, several leaders in psychedelic therapist training continue to argue that personal psychedelic experience can offer insights into the phenomenology of psychedelic states, deepen therapists’ trust in the process, and facilitate embodied therapeutic presence (Dames et al., 2024; Grof, 2008; Phelps, 2017). Despite these views, empirical research on the effects of psychedelic experiential therapist training remains almost non-existent.

There is some research evidence to suggest that psychedelic experiential therapist training may improve psychedelic therapists’ skills and expertise. The last clinical research training program to include the provision of self-experience with a classic psychedelic was the Spring Grove LSD training study conducted at the Maryland State Psychiatric Institute from 1969 to 1976. Results from a recent secondary and partial analysis of these data indicate that participants experienced personal, educational, and professional benefits from having an LSD experience (Nielson & Guss, 2018). More recently, trained therapists participated in a brief MDMA-assisted training protocol as part of their training to provide MDMA-assisted therapy through the Multidisciplinary Association for Psychedelic Studies (MAPS; NCT01404754). Results from this ‘MT-1’ study have not been published, however, according to unpublished study data shared with the authors, 86% of participants indicated that experiencing MDMA was greatly beneficial for their training, and 89.9% said that it greatly improved their qualification to conduct MDMA-assisted therapy.

Despite theoretical justifications and some research evidence in support of psychedelic experiential therapist training, the topic remains contentious. Popular press criticisms of psychedelic science have questioned whether personal use of psychedelics by research therapists could compromise scientific objectivity (Kious et al., 2023). Meanwhile, some researchers suggest personal psychedelic use may give rise to ‘psychedelic evangelism’, whereby therapists risk harming patients by overstating the potential benefits and understating the potential risks of PAT (Rosenbaum et al., 2024). Others still have expressed concern about potential social and institutional pressure to participate in psychedelic experiential therapist training (Emmerich & Humphries, 2023). These discussions further illustrate the need for empirical investigations to assess the potential risks and benefits of psychedelic experiential therapist training.

We sought to contribute to these investigations in a world-first study assessing the utility of psilocybin experiential therapist training in a sample of mental healthcare professionals who were engaged in a training to provide psilocybin-assisted therapy to individuals with Generalised Anxiety Disorder within a clinical trial (PsiGAD-1). This study, called the Psychedelic Experiential Therapist Training Study (PETT-1) sought to answer the following research questions based on the perspectives of the research therapists who participated in the psilocybin training module:

1) What are the expected professional impacts of experiencing psilocybin with support?
2) Is a psychedelic experiential therapist training module necessary and/or sufficient for the training of psychedelic therapists?
3) What are key recommendations for psychedelic experiential therapist training design and implementation?

## Method

### Trial Oversight and Study Design

PETT-1 was designed and approved as a sub-study within the PsiGAD-1 clinical trial, an investigator-initiated trial Sponsored by Monash University, approved by the Monash University Human Research Ethics Committee (Project ID 29947) and funded by Incannex Healthcare Limited. Psilocybin was provided by Usona Institute.

PETT-1 was an open label single group design wherein participants received one 25 mg dose of synthetic psilocybin with psychological support (see the *procedure* section below for further detail). The study used a qualitative design, with descriptive statistics included to facilitate a richer understanding of participant perspectives. Data collection occurred between April and November 2022.

### Conceptual Approach

A *constructionist* epistemology underpinned the study, with meaning-making understood to be iteratively constructed by participants, therapists, and researchers. The researchers took an *interpretivist* theoretical approach, whereby therapist knowledge was understood to be subjective, culturally and historically situated, and based on their interpretation of their experience (Creswell, 2003).

### Participants

Participants were 14 trial therapists who elected to participate in the study as an optional training module, following completion of a comprehensive ∼100 hour psilocybin-assisted therapy training program. Not all therapists on the PsiGAD-1 trial opted to participate. Key eligibility criteria included being a therapist on the PsiGAD-1 trial, being aged 18 to 75 years old, and agreeing to abstain from excluded psychoactive medications for the duration of the intervention. Key exclusion criteria were largely the same as for the clinical anxiety trial participants, and included being pregnant or nursing, and having a specified medical condition.

### Procedure

Research therapists were verbally informed about PETT-1 during their psychedelic therapist training. Those who expressed interest in participating were invited to complete an online screening survey which included a participant information and consent form. Participants who remained eligible following online screening assessments were invited to attend an in-person screening assessment and complete the full informed consent procedure. Participants were emailed an additional document outlining considerations associated with engaging in multiple relationships, and confidentiality measures within the therapist group.

Once enrolled in the trial, participants were engaged in numerous collaborative conversations regarding their involvement in this study, with each participant being invited to contribute their preferences and perspectives to its design and conduct. Ethical and safety considerations and measures were applied in a manner similar to the clinical participants, with some additional safeguards including establishing multiple lines of reporting, participant choice in their ‘therapists’, and strict constraints on access to the data and video of their colleagues. In line with their preferences, participants were allocated a ‘therapist’ dyad comprising one male and one female for their intervention duration. Participants had all completed the didactic component of their training prior to participating in the psilocybin training module.

The psilocybin training module comprised three sessions that emulated a truncated version of the PsiGAD-1 clinical trial that they would go on to work within:

1. *Preparation Therapy*: One 90 min therapy session conducted within the week prior to dosing, with an emphasis on psychological and practical preparation for dosing.
2. *Dosing with psychological support*: One 6-9 hr psilocybin session with a key focus on mindset, a conducive physical environment, trust, and participant-led support. An inner-directed approach was encouraged, wherein participants lay with an eye mask and headphones, and listened to a music playlist. Supportive touch for distress management was provided in accordance with participant preferences, and a benzodiazepine was available as a ‘rescue’ medication for adverse events (e.g., severe distress) that failed to respond to psychotherapeutic support.
3. *Integration therapy*: One 90 min therapy session conducted the day following dosing with emphasis on understanding and integrating key insights to sustain benefits.

### Measures and Data Collection

Quantitative study data were collected and managed using REDCap (Harris et al., 2019). Qualitative interviews were conducted by the first author, either in-person at the study site or using video conferencing software, according to participant preference. Other measures, including acute effects, were assessed and will be reported elsewhere.

### Therapist Training Survey (TTS; 3 Weeks Post-Dosing)

Adapted from the MAPS MT-1 therapist training trial, the 4-item post-intervention TTS assessed self-perceived benefits and harms associated with participation in the psilocybin training module, and the perceived importance of psilocybin experiential therapist training.

### Semi-Structured Qualitative Interviews (7-days Post-Dosing)

The interview schedule was informed by the research questions and key relevant themes identified in the literature. Participants were asked how the psilocybin experiential module impacted the way they thought about working with participants, and their opinion on a variety of associated topics such as supportive touch, multiple relationships, and risks of participation. Interviews ranged between 60-120 minutes with an average duration of 95 minutes. They were audio recorded and transcribed verbatim by a member of the research team. The interview schedule is provided in *Appendix 1*.

### Analytic Approach

Qualitative interview transcripts were checked for accuracy and imported into the qualitative data management program NVivo Plus (version 1.7.2, Lumivero 2023). Thematic analysis was used to analyse interview data in accordance with the method outlined by Braun & Clarke (2013). An initial coding framework was collaboratively developed by all authors, based on key themes identified in the literature. During analysis, initial codes were refined and new codes added to reflect new concepts in the data. The first transcript was collaboratively coded by all members of the research team, with subsequent transcripts coded by the first author in close consultation with team members. Distinctive and coherent candidate themes were identified, and the researchers returned to the dataset to ensure that all themes captured the meaning of the dataset as it related to the research questions. Short excerpts from the transcripts were selected to illustrate themes based on the spread of data therewithin. Excerpts presented in this paper have been edited to enhance readability. To ensure dependability, we kept a methodological journal where we documented the coding and theming process, and the impact of methodological choices. Member checking was completed by incorporating participant feedback on their own quotes and on a video presentation of results.

Quantitative data were analysed and visualised using R Statistical Software (v1.4.0.3; R Core Team 2020). As survey responses were mandatory, no data were missing.

Following data analysis, quantitative and qualitative results were merged for analysis, comparison, and discussion. Several quantitative items from online surveys corresponded with qualitative responses from semi-structured interviews. These results were merged at the item level to assess whether findings were triangulated and facilitate illustration.

## Results

### Sample Characteristics

We analysed data from a subset of 12 participants who continued on to provide direct therapy (as opposed to other medical or clinical support) on the PsiGAD-1 clinical trial and were thus able to participate in long-term follow-up interviews. *Table 1* presents demographic characteristics of participants in the final sample.

**Table 1.**
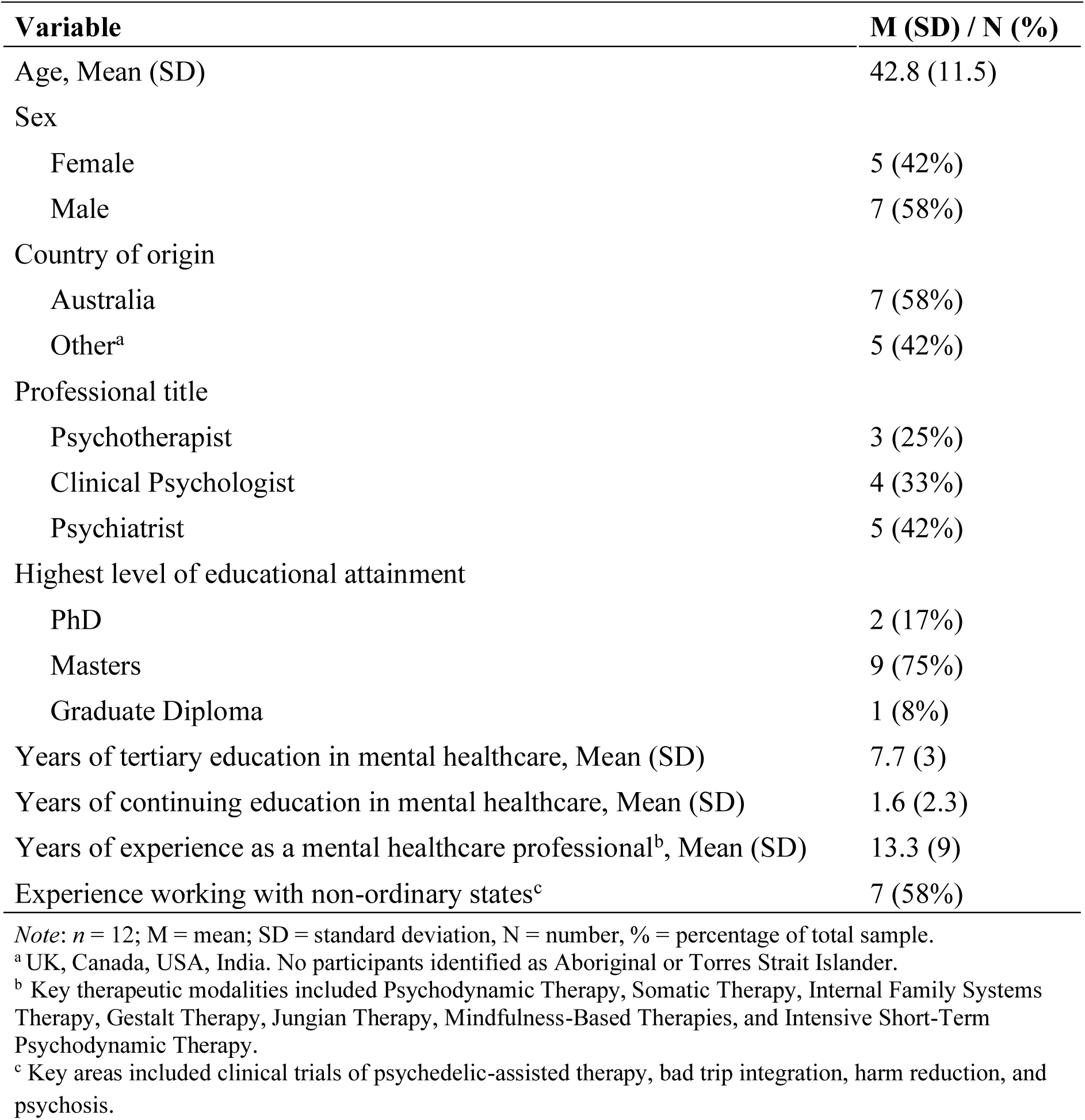
Participant demographic characteristics.

| Variable | M (SD) / N (%) |
| --- | --- |
| Age, Mean (SD) | 42.8 (11.5) |
| Sex |  |
| Female | 5 (42%) |
| Male | 7 (58%) |
| Country of origin |  |
| Australia | 7 (58%) |
| Other<sup>a</sup> | 5 (42%) |
| Professional title |  |
| Psychotherapist | 3 (25%) |
| Clinical Psychologist | 4 (33%) |
| Psychiatrist | 5 (42%) |
| Highest level of educational attainment |  |
| PhD | 2 (17%) |
| Masters | 9 (75%) |
| Graduate Diploma | 1 (8%) |
| Years of tertiary education in mental healthcare, Mean (SD) | 7.7 (3) |
| Years of continuing education in mental healthcare, Mean (SD) | 1.6 (2.3) |
| Years of experience as a mental healthcare professional<sup>b</sup>, Mean (SD) | 13.3 (9) |
| Experience working with non-ordinary states<sup>c</sup> | 7 (58%) |

*Note:* $n = 12$ ; M = mean; SD = standard deviation, N = number, % = percentage of total sample.
<sup>a</sup> UK, Canada, USA, India. No participants identified as Aboriginal or Torres Strait Islander.
<sup>b</sup> Key therapeutic modalities included Psychodynamic Therapy, Somatic Therapy, Internal Family Systems Therapy, Gestalt Therapy, Jungian Therapy, Mindfulness-Based Therapies, and Intensive Short-Term Psychodynamic Therapy.
<sup>c</sup> Key areas included clinical trials of psychedelic-assisted therapy, bad trip integration, harm reduction, and psychosis.

### Qualitative Themes

*Table 2* presents three main themes that were constructed, and the sub themes that characterised each. Themes are illustrated in greater detail below.

**Table 2.** Qualitative themes and subthemes.

| Theme | Subtheme |
| --- | --- |
| Professional benefits of psilocybin experiential therapist training | Experiencing the subjective effects of psilocybin in a therapeutic context enhanced core therapist qualities<br>Gaining an embodied understanding of key therapeutic principles enhanced therapeutic skill |
| Speculated risks of psychedelic experiential therapist training | Therapists may overgeneralise their psychedelic experience and provide care based on a restricted frame of reference<br>Therapists may experience a period of personal destabilisation |
| Recommendations for psychedelic experiential therapist training design and implementation | Psychedelic experiential therapist training is necessary but not sufficient for the provision of the highest quality practice in psychedelic therapy<br>Professional preparation and integration frameworks are key to psychedelic experiential therapist training outcomes |

**Theme 1: Professional Benefits of Psilocybin-Assisted Training**

*Figure 1* presents participant ratings of the extent to which the psilocybin training module felt professionally beneficial. Results indicate that participants perceived their psilocybin experience to have a substantial positive impact on their capacity to provide PAT.

**Figure 1.**
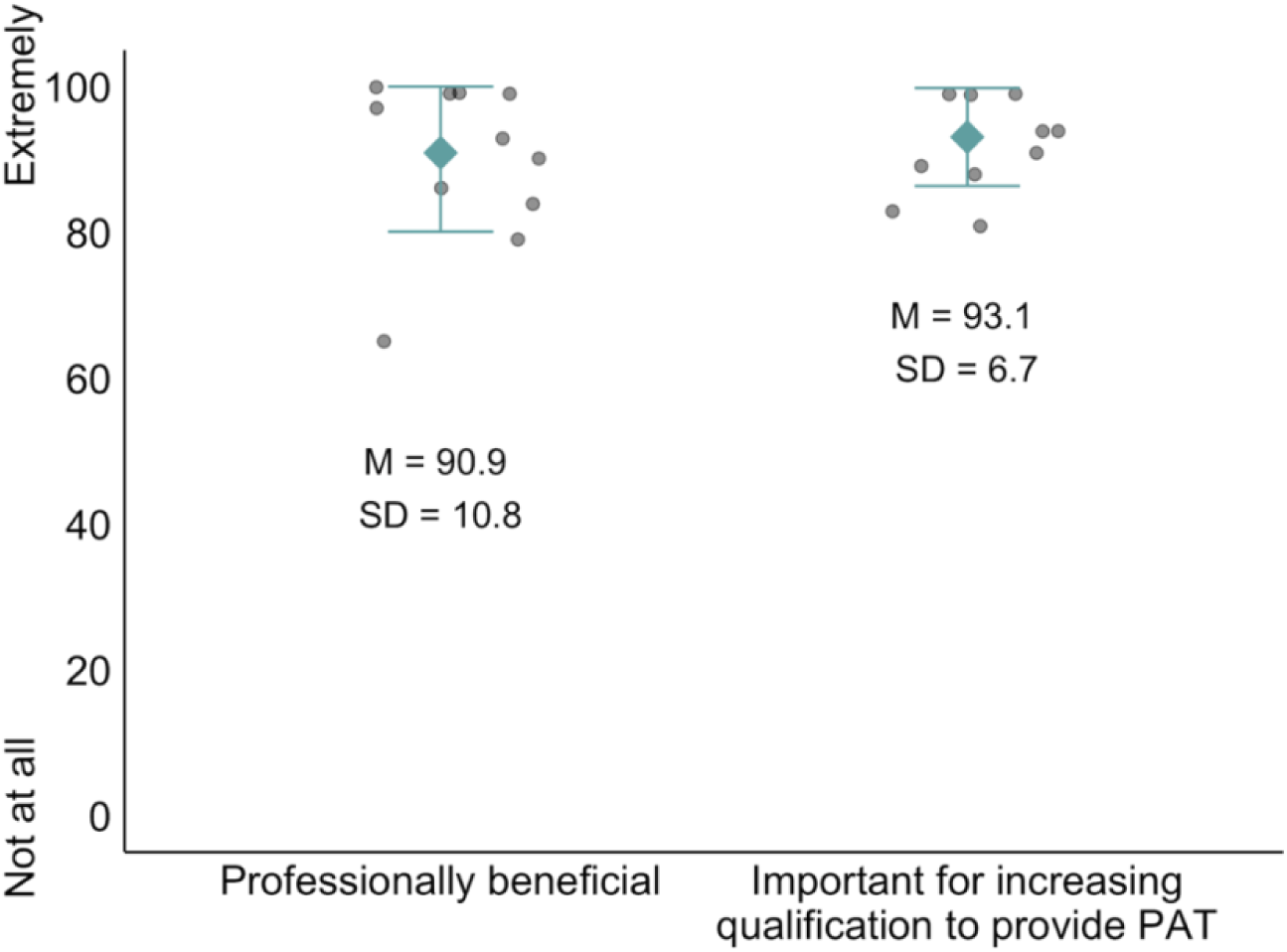
Participant ratings of the professional benefits of psilocybin-assisted training. *Note*: M = mean, SD = standard deviation, *n* = 12, PAT = psilocybin-assisted therapy; the mean and standard deviation of item responses are depicted in green; raw datapoints are represented by grey circles.

As illustrated below, there are two key ways in which participants felt the psilocybin-assisted training was professionally beneficial. First, it fostered core therapist qualities. Second, it enhanced therapeutic skill.

### 1.1 Experiencing the Subjective Effects of Psilocybin in a Therapeutic Context Enhanced Core Therapist Qualities

*Figure 2* presents participant ratings from the Therapist Training Survey of the impact of the psilocybin experiential module on various therapist qualities.

**Figure 2.**
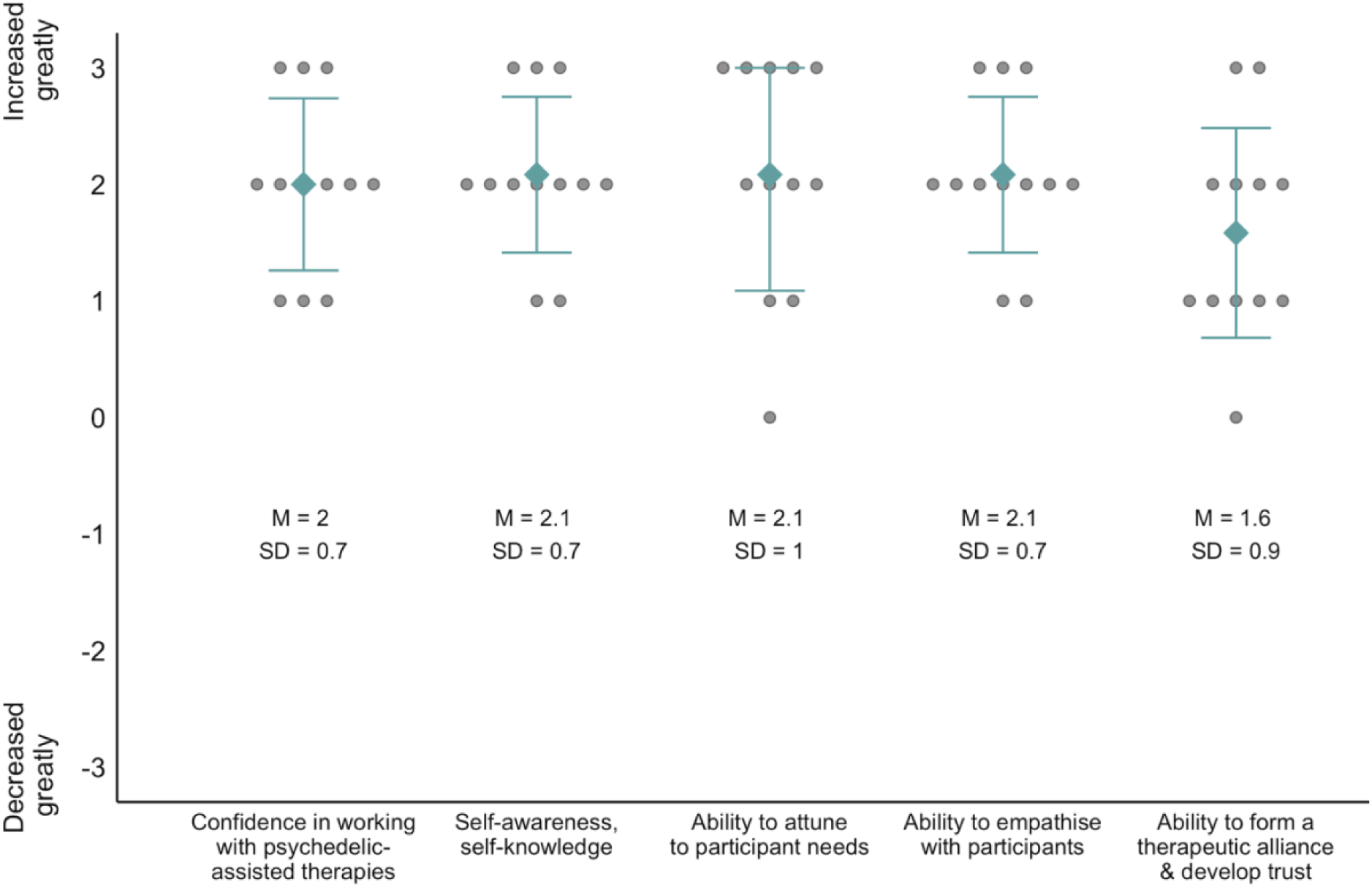
Participant ratings of the impact of psilocybin-assisted training on therapist characteristics. *Note*: M = mean, SD = standard deviation; 0 = neither increased nor decreased, *n* = 12; the mean and standard deviation of item responses are depicted in green; raw datapoints are represented by grey circles.

Results indicate that participants felt the psilocybin training module positively impacted their confidence and capacity for self-awareness, attunement, empathy, and facilitating trust and therapeutic alliance. As illustrated below, qualitative results triangulated these findings and illuminated the important contribution of first-hand experience with both psilocybin’s subjective effects and the therapeutic context to these increases.

#### 1.1.1 Experiencing Subjective Effects of Psilocybin

All participants emphasised the translational value of experiencing common psychedelic phenomena first-hand for their work with clinical participants. In particular, phenomena that cannot be reliably catalysed to the same extent via other means such as ineffability, spiritual experiences, veridicality, sudden ontological shifts, and a loss of control:

> *“I am not of the school of thought that says I need to have had an experience to be able to really well support someone else through that. I do think this experience stands apart. In all those other experiences, you’re an ego identity navigating a challenge. Whereas in this experience, you’re no longer that ego identity. And so it’s quite unlike any other ordeal a person will go through.” - P2*

All participants reported an increase in cognitive empathy (i.e., intellectual understanding of others’ experiences) for clinical participants as a result of personally encountering these psychedelic phenomena. They anticipated that this would increase their capacity to accurately interpret and work therapeutically with participant experiences during dosing and integration:

> *“It takes someone with a sophisticated grasp of language to convey [common psychedelic phenomena], but even then, there needs to be a recipient to that language that can subjectively understand and translate that meaning.” - P8*

All participant accounts implicitly evidenced increased emotional empathy (i.e., understanding and resonating with others’ affective states) for clinical participants following their own psilocybin-catalysed emotional and somatic experiences. Participants projected that increased emotional empathy would positively impact work with participants by increasing compassion, common humanity, emotional connection and attuned empathic support (e.g., “*Having had the experience makes you more able to feel into what is needed through moments of suffering or great love.” - P8*).

All participants described increased confidence in their ability to safely and effectively provide PAT (self-efficacy) following the psilocybin training module. Contributing factors to these increases included feeling enhanced credibility and increased trust towards the study drug. Responses implied that these increases would help to facilitate trust and engender safety with participants:

> *“I think it’s trust and confidence. But not theoretical trust and confidence; embodied trust and confidence. Because in that state, the state of the therapist is extremely contagious…If the participant is anxious, and the therapist is in part informed by having been through this, is in a very open calm state, that embodied trust in what’s happening will pass to the participant*.” - *P2*

#### 1.1.2 Experiencing the Therapeutic Context

All participants emphasised the contribution of the therapeutic context to learning outcomes (e.g., “*Having a psychedelic experience wandering through a forest is very different from having a psychedelic experience where you’re going 95% inwards with music, with people witnessing”* - P3). In the dosing context, most participants reported increased understanding and empathy around the interaction between the study drug and the physical environment (e.g., using a shared restroom) within the framework of the trial protocol (e.g., internally focussed approach with eye shades and headphones). Several participants also demonstrated increased empathy for the challenges associated with transitioning from the dosing environment to the home environment in a vulnerable state. This increased a sense of importance around helping clinical participants adequately prepare emotionally, physically, and practically for such transitions (e.g., *“I came out being much more mindful of doing whatever I can in prep and dosing to help participants navigate those rapid transitions between contexts.” - P2*).

Most participants emphasised the importance of experiencing the relational context of PAT. For example, the magnitude of vulnerability required of participants, and the level of trust, safety, and therapeutic rapport needed to support this vulnerability. Based on their experience, participants reported acquiring an embodied understanding of the importance of therapist qualities that support these factors such as respect for ethical boundaries, comfort with intimacy, unconditional positive regard, and authenticity (e.g., “*The honesty and authenticity of those individuals comes through and is felt within” - P1*).

Several participants posited that the ability to disclose their psilocybin experience, where therapeutically useful, would facilitate trust and safety with clinical participants (e.g., *“I can confidently say ‘Yes, I’ve done this, I’ve done it right here’. And that closeness of experience, I think, feels really supportive”* - P2). In two cases, this learning was based on personal experience during the psilocybin training module (e.g., “*Now I get self-disclosure differently because my therapist used it and it landed in a really important way."* - P7).

Most participants spoke to the value of experiencing the co-therapy model as a participant. Here, one key learning was how important the quality of the relationship between co-therapists was throughout treatment for creating a sense of relational safety (e.g.: *“Any dynamic between the therapists in the dyad will be amplified. If there’s a sense of relational ease, and deep trust and understanding between the therapists, that’s very supportive of the participants’ process.”*- P2). A second key learning was based on experiencing increased transference due to the presence of two therapists, and amplification of interrelationships during dosing (e.g., *“I perceived my therapists in a very relational way. I felt like my parents were looking on*” - P11). Based on this experience, participants demonstrated greater sensitivity towards clinical participants’ potential transference and endorsed the promise of the co-therapy model for facilitating a corrective experience. In some cases, such experiences resulted in a shift in philosophical perspective on how therapeutic approaches are applied in dosing. For example, some participants came to value a more flexible approach that balances inner-directed (described below) and relational processes.

### 1.2 Gaining an Embodied Understanding of Key Therapeutic Principles Enhanced Therapeutic Skill

All participants reported gaining an embodied (i.e., conceptual knowledge derived from subjective sensorimotor experience) understanding of key principles of PAT which they previously understood theoretically from the didactic component of their training. They projected this would increase their competence to provide PAT via increased understanding of why, how, and when to implement therapeutic techniques. Key principles cited are detailed below, and include an inner-directed approach, supportive touch, and how to support preparation and integration.

#### 1.2.1 Inner-directed Approach

The therapeutic model presented in participants’ training emphasised allowing somatic, affective, and psychological experiences to unfold during psilocybin dosing, framing these experiences as important sources of insight and therapeutic processing. Many participants indicated that experiencing a this so-called ‘inner-directed’ approach fostered a deeper understanding of the therapeutic process, and highlighted the importance of ‘unlearning’ the more directive approaches they were familiar with from delivering conventional talk therapy:

> *“The main thing it did was help me trust individuals’ internal healing to actually happen, and to stay out of the way even more than what I have thought was appropriate. I really do feel like it helped me fully trust the process [to] unfold, be present, and really do not try to direct or influence, because it’s like there’s this hurricane coming and you’re shouting in the wind…there’s no point.” - P6*

Practically speaking, participants reported gaining clarity around the importance of reducing participants’ external cognitive load (e.g. minimising questions and trial surveys, refraining from guiding or interpreting individuals’ experience) and taking care of physical and emotional needs (e.g., “*The feeling of just being cared for in that space is extraordinary*” - P6). Experiencing the impact of such non-directive support, driven by therapist qualities such as presence, compassion, attunement, and genuineness, enhanced the rationale for applying this approach with clinical participants:

> *“They were just there, present, listening, caring, loving. That’s all I needed*. *That’s what I hope I can give to somebody else” -P1*

> “*You’re sitting there just being present for the person. But when I’ve done it so far, I haven’t trusted that the client has really felt it. Well, now I know that the client can feel it quite intensely, cause I did. That helps me to understand the importance of just being there.” -P11*

#### 1.2.2 Supportive Touch

Most participants reported experiencing supportive touch as a useful tool for grounding and distress management during peak drug effects when their cognitive-linguistic capacity was limited. These participants tended to describe resultant increases in their understanding around the rationale, sense of importance, and capacity to deliver supportive touch:

> *“I was someone who prior to that experience was very much like…’do we really need to practise therapeutic touch at all?’…But in my experience it was huge and important… it was often the thing that would help me get a bit of a breakthrough.” -P7*

Two participants however, felt ambivalent about the net therapeutic impact of supportive touch and pondered whether their therapists provided it in response to their own needs. One such participant recommended a cautionary approach to providing supportive touch in PAT:

> *“The risks are substantial…and go beyond merely the therapist’s credibility, reputation, and registration to this whole undertaking of psychedelic therapy…The touch was lovely but was it essential to my experience? I’m still not really convinced.” - P8*

Irrespective of their experience of supportive touch, all participants agreed on the importance of collaboratively developing, practising, and following clear guidelines around touch in PAT. This was born of experiencing a deep sense of trust, respect, and safety from an approach that emphasised clear detailed communication and respect for boundaries.

#### 1.2.3 Preparation and Integration

Several participants expressed a clearer understanding of the principles and processes of effective preparation and integration based on their own experience. Key learnings centred on how to adequately prepare participants psychologically for the psychedelic experience, how to support participants’ significant others so they can in turn support participants, how psychedelic-catalysed insights unfold over time, and methods of integrating and staying in touch with the psychedelic experience.

**Theme 2: Speculated Risks of Psychedelic Experiential Therapist Training**

Quantitative and qualitative responses indicated that participants perceived no personal or professional harms from participation. However, they speculated two ways in which psychedelic experiential therapist training could potentially negatively impact therapists’ capacity to provide PAT: by restricting their frame of reference or contributing to a period of personal destabilisation.

### 2.1 Therapists May Overgeneralise their Psychedelic Experience and Provide Care Based on a Restricted Frame of Reference

Most participants were concerned about the risk of therapists projecting their own experience onto those of clients in a manner that narrows interpretative range, reduces attunement, and undermines alliance. This was explicitly addressed by two participants who speculated that psychedelic-naive trainees in particular could lack critical appraisal of what they encounter during their psychedelic experience:

> *“If this is someone’s first, or first of a handful of peak experiences, there’s an initial relationship with what’s been encountered which is often overly simplistic and involves reification and concretisation of what has been encountered…that relationship with what has been encountered I think needs to be navigated with care.” - P2*

Resultant failure of therapists to listen and respond effectively may render their clients less likely to express or feel things that sit outside of their therapists’ frame of reference.

### 2.2 Therapists May Experience a Period of Personal Destabilisation

Several participants speculated that trainees’ psychedelic experience could elicit challenging material that may destabilise them for a period. If insufficiently integrated, this could decrease therapists’ trust in PAT and reduce their capacity to engender safety and support clients through challenging experiences (e.g., *“A bad trip may trigger aversion, anxieties, and fears in [therapists] and interfere with their ability to connect”* - P11). Participants recommended additional integration sessions and/or personal therapy following challenging experiences to support trainees’ wellbeing and ensure that impactful material is sufficiently integrated prior to providing PAT (e.g., “*Having that personal and professional space to discuss it would be very important because even a bad trip tells you something about yourself”* - P11).

**Theme 3: Recommendations for Psychedelic Experiential Therapist Training Design and Implementation**

### 3.1 Psychedelic Experiential Therapist Training is Necessary but not Sufficient for the Provision of the Highest Quality Practice in Psychedelic-Assisted Therapy

Participants unanimously endorsed that direct personal experience of psychedelics in a therapeutic context is a necessary but not sufficient prerequisite for trained healthcare professionals to provide the highest quality of care in PAT. These results are triangulated and extended by qualitative findings:

#### 3.1.1 Necessity

Participant views varied on whether direct experience with psychedelics should be a prerequisite for becoming a psychedelic therapist. However, all agreed that it is necessary for providing the highest quality of care in PAT (e.g., *"I was [providing psychedelic] therapy before I’d had that experience and I think I was still doing a good job…but I think the quality of what you can offer really shifts."* - P9). This was chiefly based on benefits they gained from their own psilocybin experiential therapist training that augmented the level of detail and quality of care in their work. Unique benefits included embodied knowledge and greater self-knowledge (e.g., *“It’s such a helpful way of understanding your own psyche. This work is really all about helping people understand their own psyches, and because of transference you have to understand your own psyche to help them understand theirs” -* P4). Therapists felt these learnings could not be catalysed to the same extent via other means:

> *“I think it’s a fundamentally different type of care you can provide, if you have, versus haven’t, had the experience yourself. I wouldn’t learn to play basketball from someone who had never shot a hoop.” - P7*

Participants recognised a distinction between choosing not to, or being unable to, participate in psychedelic experiential therapist training. They suggested it may be useful to collaboratively assess whether reservations about participation point to professional domains that would benefit from further development prior to providing psychedelic therapy (e.g., low openness to experience, unwillingness to engage in meaningful personal development, personal defences). In cases where psychedelic experiential therapist training is inaccessible or unadvisable (e.g., due to contraindications), participants recommended participating in activities associated with flow states, peak/challenging experiences, and non-ordinary states as alternative means of knowledge and skill development.

#### 3.1.2 Sufficiency

Participants agreed that psychedelic experiential therapist training alone is not sufficient for the provision of the highest quality of care in PAT. Rather, knowledge and skill from psychotherapeutic training are foundational to the provision of safe and effective PAT. Here, key areas include: therapeutic skill and theoretical knowledge relevant to clinical populations; ethical conduct; risk management; recognising and working with transference and countertransference; and self-awareness/reflective capacity.

Participants also highlighted the importance of embedding psychedelic experiential therapist training within comprehensive PAT training programs. Indeed, their interview responses demonstrated that metacognitive awareness of principles and process of PAT learned in non-drug-assisted components of their PAT training informed what they learned during the experiential component.

Taken together, participant responses highlight the importance of combining therapeutic knowledge and expertise with psychedelic experiential therapist training to optimise therapeutic outcomes:

> *“There are these people who do deep sea engineering…Taking a psychedelic is like being able to scuba dive, and being an engineer is like being able to work with what you encountered down there. And being a psychedelic therapist is putting the two together. So [without therapeutic training] it’s like sending a scuba diver down to deal with the oil rig, but they haven’t got an engineering degree, so they don’t have a clue what to do once they’re down there.” - P2*

### 3.2 Professional Preparation and Integration Frameworks are Key to Psychedelic Experiential Therapist Training Outcomes

Participant responses indicate that the professional impact of psychedelic experiential therapist training can be optimised via professional preparation and integration frameworks. Elements of these frameworks include: framing the psychedelic experience in a way that promotes humility and minimises overgeneralisation; setting a professional intention for dosing; providing detailed reflective prompts to assist trainees to interpret their experience in the context of their work as psychedelic therapists; where applicable, creating a forum for trainees to provide feedback to other trainees acting as their therapist(s); and ensuring meaningful engagement in good individual and group supervision with exposure to multiple experiences and perspectives. The implementation of these elements ought to align with the timing of the experiential learning process:

> *“Although we report these experiences in a narrative or cognitive way, so much of what has happened for us is somatic or affective. And so having a kind of an intense challenging philosophical back and forth around narrative and belief could be very emotionally and systemically jarring. So ideally, you’d have your self-experience, you’d have three or four weeks, then you’d have supervision, before people go and sit with participants” - P2*

This is particularly relevant in light of the fact that participants appeared to move through the process of experiential learning at different rates depending on several factors, including their baseline level of experience with non-ordinary states of consciousness.

Further to professional frameworks, several participants spoke to the importance of embedding psychedelic experiential therapist training in ongoing personal therapy to mitigate potential risks and enhance professionally relevant learnings.

## Discussion

### Psilocybin Experiential Therapist Training: Impacts on Training Outcomes and Implications

Results indicate that direct personal experience with psychedelics in a therapeutic context may meaningfully increase the quality of care that therapists can provide in PAT. The degree of reported impact in the current sample is notable considering it comprised mental healthcare professionals with substantial training and experience (including working with non-ordinary states of consciousness) who possessed strong core therapist qualities prior to participation. Results align with previous research findings (Nielson, 2021) and provide novel support for the inclusion of psychedelic experiential therapist training within PAT therapist training programs.

Participants demonstrated increases in cognitive and emotional empathy from direct experience of the same drug in the same therapeutic context as clinical participants. Participants felt that key experiences underpinning increases in empathy could not be catalysed to the same extent via other means. Empathy is understood as a key change mechanism across multiple psychotherapeutic modalities (Watson et al., 2013). In the current context, participants projected that increased empathy would enhance their capacity to understand and respond effectively to clinical participants.

Increases in empathy catalysed increases in other therapist qualities including attunement, compassion, common humanity, authenticity, and trust facilitation. Alongside empathy, these qualities are foundational to a therapist’s capacity to build a strong therapeutic alliance and engender safety (Aponte, 2022; Gilbert, 2020; McClintock et al., 2018; Nienhuis et al., 2018; Prusiński, 2022; Wampold, 2015). Importantly, emerging evidence suggests that the strength of the therapeutic alliance influences treatment outcome in PAT both directly and via increases in emotional breakthrough and mystical-type experience during peak drug effects (Murphy et al., 2021).

Results also suggest that psychedelic experiential therapist training may increase two domains of therapist well-being that impact therapeutic function (Moe & Thimm, 2021). First, participants reported increased professional self-efficacy, which has been found to be positively associated with well-being and work performance in healthcare workers (Bali-Mahomed et al., 2022; Ghani et al., 2024). Second, participants reported feeling greater empathy and attunement with clinical participants, such that compassionate and caring responses are more likely to be genuine, with little dissonance between their felt and expressed emotion. In literature examining how healthcare professionals manage and express emotion to provide compassionate care (referred to as ‘emotional labour’), this kind of congruence would align with a concept called ‘deep acting’. This stands in contrast to instances where a therapist may need to exert more effort because of a higher level of dissonance between their felt and expressed emotion. For example, by suppressing mood-incongruent emotions or trying to empathise with experiences outside their understanding (referred to as ‘surface acting’). Though limited, the research evidence in this area suggests that for healthcare workers, deep (as opposed to surface) acting, protects against burnout and promotes wellbeing and work performance (Hülsheger & Schewe, 2011; Mesmer-Magnus et al., 2012; Yeh et al., 2020).

Participant responses indicate that experiential learning facilitated embodied knowledge of key therapeutic principals which in turn increased understanding around why, how, and when to implement therapeutic techniques. Such embodied knowledge is particularly relevant for work with psilocybin-assisted therapy, wherein therapists support psychological change processes involving the navigation of somatic, affective, and/or psychodynamic experiences. Findings support extant theoretical approaches to training professionals in the provision of other psychotherapeutic modalities involving changes to waking consciousness such as psychodynamic therapy and mindfulness-based therapies, where experiential learning is considered an essential training component (Crane et al., 2012; Feinstein et al., 2015).

More broadly, the current study illustrates how psychedelic experiential therapist training can generate critical discussion around understudied aspects of PAT that have hitherto been based on received wisdom, thereby generating new questions and new knowledge (e.g., the role and importance of supportive touch, the optimal balance between a relational vs inner-directed approach in dosing). Likewise, first-hand experience of PAT by individuals with awareness of therapeutic frameworks can help to improve understanding around mechanistic processes and how best to support them therapeutically. In this instance, participant feedback directly influenced the trial protocol (e.g., some dose-day surveys were eliminated in support of an inner-directed approach) and personal experiences informed group discussions that shaped the collective therapeutic approach. Furthermore, participants can carry this knowledge into clinical service delivery.

### Optimising Outcomes and Mitigating Potential Risks

Participants agreed that experiential learning consolidated an embodied knowledge of concepts they previously understood intellectually from didactic training. Further, participants indicated that prior therapeutic knowledge, skill, and core qualities were essential to both the way therapists interpreted their psychedelic experience, and their capacity to provide high quality care in PAT. This suggests that psychedelic experiential therapist training may be useful when delivered within comprehensive psychedelic therapist training programs to mental healthcare professionals selected for relevant knowledge, skills, and core qualities.

Participants recommended that the benefits of psychedelic experiential therapist training could be augmented by ensuring that the experiential component is delivered within appropriate reflexive frameworks. Participants recommended that such frameworks should: encourage meaningful engagement in individual and group supervision; facilitate critical appraisal of the psychedelic experience; promote both self-efficacy and humility; incorporate structured professional reflective practice; include feedback mechanisms; provide forums for critical group discussion and exposure to diverse experiences of psychedelic experiential therapist training; encourage personal therapy; and provide access to subsequent psychedelic experiential therapist training sessions to increase the range of experiences trainees can draw on in their work with clients. Such frameworks can augment professionally applicable learnings and mitigate potential risks such as frame restriction and destabilisation.

Finally, as participants appeared to move through the process of experiential learning at different rates, professional frameworks should be developed with sensitivity to individual differences in processing time and level of support required. Models that capture experiential learning processes (e.g., Kolb & Fry, 1974) can be explored as a means of guiding these frameworks.

### Limitations & Future Directions

This study had a small sample size with no comparator group or validated measures. As such, while results indicate utility for psychedelic experiential therapist training, they are preliminary in nature. The sample itself comprised experienced mental healthcare professionals attuned to a range of clinical and ethical issues specific to PAT and demonstrated strong self-reflective capacity. These qualities were selected for and reinforced by the trial design, which included a rigorous therapist selection process, and structured opportunities for individual and group reflection. This limits generalisability to other contexts. However, responses within this study highlight the value of selecting individuals with appropriate training and expertise to participate in psychedelic experiential therapist training programs. Results should be validated and extended in larger studies with comparator groups, comprising samples that are more diverse with respect to professional qualifications and experience. Such studies should include validated pre- and post-measures that control for baseline therapeutic capacity.

Therapists who declined participation were not interviewed. As such, only the views of those who were willing and able to participate in psychedelic experiential therapist training are reflected in the results. This sample may have harboured biased expectations about the utility of psychedelic experiential therapist training which inflated the benefits they perceived from participation. This is particularly relevant considering all measures included were self-report. Further research is needed to facilitate an understanding of a greater variety of perspectives and incorporate external ratings.

Qualitative findings suggest at least three valuable avenues for future research. First, participant responses provided insight into what constitutes key psychedelic therapist competencies and qualities. To optimise and assess training outcomes, future research could incorporate this information into a framework of psychedelic therapist competencies that can be empirically tested and then deployed for assessing individual competencies and designing and assessing training programs. Second, participant responses one-week post-dosing illustrated individual differences in the pace of personal and professional integration. It may be useful to characterise the various stages of experiential learning in psychedelic experiential therapist training so that such differences can be more readily identified, and supportive frameworks can be tailored accordingly. Finally, as mentioned above, results indicate that psilocybin-assisted training may change the way in which therapists provide emotional labour. However, emotional labour is a relatively understudied construct in healthcare and the relationship between emotional labour and PAT is yet to be explored. Further research into this area would contribute to theoretical understandings of how psilocybin-assisted training influences how therapists provide compassionate care, and how this impacts both therapist wellbeing and therapeutic function.

### Concluding Remarks

Results from this world-first study indicate that psilocybin-assisted therapist training may meaningfully increase therapists’ capacity to provide PAT. Despite the study’s limitations, it was successful in characterising the potential benefits and risks of psychedelic experiential therapist training and generating a set of recommendations for its design and implementation. Findings suggest that psychedelic experiential therapist training may be uniquely valuable for developing certain psychedelic therapist capacities, and if so, may play a meaningful role in the delivery of safe and effective psychedelic-assisted therapy.

### CRediT Authorship Contribution Statement

**Georgia Ioakimidis-MacDougall** Methodology, project support, led formal analysis, investigation, writing original draft, reviewing and editing manuscript, visualisation, project administration. **Paul Liknaitzky**: Funding acquisition, conceptualisation, methodology, project management, assisted formal analysis, investigation, resources, reviewing and editing manuscript, supervision. **John Gardner**: Mentored and assisted formal analysis, reviewing and editing manuscript, supervision.

## Data Availability

Due to the sensitive and potentially identifiable nature of the qualitative data, they have not been made publicly available.

## Acknowledgments

This research was funded by Incannex Healthcare Limited and Monash University, with Psilocybin provided by Usona Institute. Thank-you to Rick Doblin, Shannon Carlin, and the Multidisciplinary Association for Psychedelic Studies for their support. Thank-you to William Richards for his mentorship, and the participants for their time and expertise. Thank-you to Joshua Kugel for proofreading the manuscript, Murat Yücel for his project mentorship, and Rachel Ham, Hannah Bushell, and the Clinical Psychedelic Lab team at Monash University for their assistance.

## Appendix 1: Semi-Structured Qualitative Interview Schedule

- Did you have any experiences or insights during the psilocybin training module that impacted the way you think about working with participants?
- As you continue to integrate your experiences and insights from the psilocybin training module, what processes or supports do you see being important for helping you to apply this knowledge to your work with participants?
- Are there any specific learnings relevant to providing psychedelic therapy that you think could only be occasioned by self-experience with psychedelics, and not via other means (e.g., meditation, self-reflection, non-drug self-experience)?
- How has your participation in the psilocybin training module impacted your working relationship with [therapist A] and [therapist B]?
- What was your experience of therapeutic touch during the psychedelic experience?
- Are there any ways in which you can see the psilocybin training module having a negative impact on therapists’ capacity to provide psychedelic therapy?
- Do you see an ongoing role for self-experience in your professional development as a psychedelic therapist?
- In what ways could the psilocybin training module be improved to better support you or your learning?
- Is there anything else that you would like to share?

## References

Aponte, H. J. (2022). The soul of therapy: The therapist’s use of self in the therapeutic relationship. Contemporary Family Therapy, 44(2), 136–143. 10.1007/s10591-021-09614-5

Bali-Mahomed, N. J., Ku-Johari, K. S., Mahmud, M. I., Amat, S., & Saadon, S. (2022). Psychological Well-Being of School Counsellors Model. European Journal of Educational Research, 11(2), 621–638. 10.12973/er-jer.11.2.621

Braun, V., & Clarke, V. (2013). Successful Qualitative Research: A Practical Guide for Beginners. SAGE.

Carhart-Harris, R., Giribaldi, B., Watts, R., Baker-Jones, M., Murphy-Beiner, A., Murphy, R., Martell, J., Blemings, A., Erritzoe, D., & Nutt, D. J. (2021). Trial of Psilocybin versus Escitalopram for Depression. New England Journal of Medicine, 384(15), 1402–1411. 10.1056/NEJMoa2032994

ClinicalTrials.gov. (2024, 20011). Psychological effects of Methylenedioxymethamphetamine (MDMA) when administered to healthy volunteers (MT-1). Multidisciplinary Association for Psychedelic Studies. https://clinicaltrials.gov/study/NCT01404754

Crane, C., Winder, R., Hargus, E., Amarasinghe, M., & Barnhofer, T. (2012). Effects of mindfulness-based cognitive therapy on specificity of life goals. Cognitive Therapy and Research, 36(3), 182–189.

Creswell, J. W. (2003). Research design: Qualitative, quantitative, and mixed method approaches (2nd ed). Sage Publications.

Dames, S., Watler, C., Kryskow, P., Allard, P., Gagnon, M., Taylor, W., & Tsang, V. W. L. (2024). Psychedelic-Assisted Therapy Training: An Argument in Support of Firsthand Experience of Nonordinary States of Consciousness in the Development of Competence. Psychedelic Medicine, 2(3), 130–137. 10.1089/psymed.2023.0004

Emmerich, N., & Humphries, B. (2023). Is the Requirement for First-Person Experience of Psychedelic Drugs a Justified Component of a Psychedelic Therapist’s Training? Cambridge Quarterly of Healthcare Ethics, 1–10. 10.1017/S0963180123000099

Feinstein, R. E., Huhn, R., & Yager, J. (2015). Apprenticeship Model of Psychotherapy Training and Supervision: Utilizing Six Tools of Experiential Learning. Academic Psychiatry, 39(5), 585–589. 10.1007/s40596-015-0280-6

George, J. R., Michaels, T. I., Sevelius, J., & Williams, M. T. (2019). The psychedelic renaissance and the limitations of a white-dominant medical framework: A call for indigenous and ethnic minority inclusion. Journal of Psychedelic Studies, 4(1), 4–15. 10.1556/2054.2019.015

Ghani, U., Usman, M., Cheng, J., Mehmood, Q., & Shao, X. (2024). Does Professional Self-Efficacy Provide a Shield in Troubling Situations? Evidence of Performance and Thriving Through Perceived Strength Use. Sage Open, 14(2), 21582440241252507. 10.1177/21582440241252507

Gilbert, P. (2020). Compassion: From Its Evolution to a Psychotherapy. Frontiers in Psychology, 11. 10.3389/fpsyg.2020.586161

Griffiths, R. R., Johnson, M. W., Carducci, M. A., Umbricht, A., Richards, W. A., Richards, B. D., Cosimano, M. P., & Klinedinst, M. A. (2016). Psilocybin produces substantial and sustained decreases in depression and anxiety in patients with life-threatening cancer: A randomized double-blind trial. *Journal of Psychopharmacology (Oxford*, England), 30(12), 1181–1197. 10.1177/0269881116675513

Grof, S. (with Hoffman, A.). (2008). LSD Psychotherapy: The Healing Potential of Psychedelic Medicine (4th ed.). Multidisciplinary Association for Psychedelic Studies.

Halberstadt, N. (2014, July 17). MDMA-Assisted Therapy: A View from Both Sides of the Couch. Multidisciplinary Association for Psychedelic Studies - MAPS. https://maps.org/news/bulletin/mdma-assisted-psychotherapy-a-view-from-both-sides-of-the-couch/

Harris, P. A., Taylor, R., Minor, B. L., Elliott, V., Fernandez, M., O’Neal, L., McLeod, L., Delacqua, G., Delacqua, F., Kirby, J., & Duda, S. N. (2019). The REDCap consortium: Building an international community of software platform partners. Journal of Biomedical Informatics, 95, 103208. 10.1016/j.jbi.2019.103208

Hoffman, A. (1980). LSD: My problem child. McGraw-Hill Book Company.

Hülsheger, U. R., & Schewe, A. F. (2011). On the costs and benefits of emotional labor: A meta-analysis of three decades of research. Journal of Occupational Health Psychology, 16(3), 361–389. 10.1037/a0022876

Johnson, M. W., Garcia-Romeu, A., & Griffiths, R. R. (2017). Long-term follow-up of psilocybin-facilitated smoking cessation. The American Journal of Drug and Alcohol Abuse, 43(1), 55–60. 10.3109/00952990.2016.1170135

Kious, B., Schwartz, Z., & Lewis, B. (2023). Should we be leery of being Leary? Concerns about psychedelic use by psychedelic researchers. Journal of Psychopharmacology, 37(1), 45–48. 10.1177/02698811221133461

Kolb, D., & Fry, R. (1974). Toward an Applied Theory of Experiential Learning. Wiley.

Mangini, M. (1998). Treatment of Alcoholism Using Psychedelic Drugs: A Review of the Program of Research. Journal of Psychoactive Drugs, 30(4), 381–418. 10.1080/02791072.1998.10399714

McClintock, A. S., Anderson, T., Patterson, C. L., & Wing, E. H. (2018). Early psychotherapeutic empathy, alliance, and client outcome: Preliminary evidence of indirect effects. Journal of Clinical Psychology, 74(6), 839–848. 10.1002/jclp.22568

McLaughlin, S. C., & Grof, S. (1976). Realms of the Human Unconscious: Observations from LSD Research. Journal for the Scientific Study of Religion, 15(4), 376. 10.2307/1385643

Mesmer-Magnus, J. R., DeChurch, L. A., & Wax, A. (2012). Moving emotional labor beyond surface and deep acting: A discordance–congruence perspective. Organizational Psychology Review, 2(1), 6–53. 10.1177/2041386611417746

Moe, F. D., & Thimm, J. (2021). Personal therapy and the personal therapist. Nordic Psychology, 73(1), 3–28. 10.1080/19012276.2020.1762713

Murphy, R., Kettner, H., Zeifman, R., Giribaldi, B., Kartner, L., Martell, J., Read, T., Murphy-Beiner, A., Baker-Jones, M., Nutt, D., Erritzoe, D., Watts, R., & Carhart-Harris, R. (2021). Therapeutic Alliance and Rapport Modulate Responses to Psilocybin Assisted Therapy for Depression. Frontiers in Pharmacology, 12, 788155. 10.3389/fphar.2021.788155

Nielson, E. M. (2021). Psychedelics as a Training Experience for Psychedelic Therapists: Drawing on History to Inform Current Practice. Journal of Humanistic Psychology, 28(4), 618–634. 10.1177/00221678211021204

Nielson, E. M., & Guss, J. (2018). The influence of therapists’ first-hand experience with psychedelics on psychedelic-assisted psychotherapy research and therapist training. Journal of Psychedelic Studies, 2(2), 64–73. 10.1556/2054.2018.009

Nienhuis, J. B., Owen, J., Valentine, J. C., Winkeljohn Black, S., Halford, T. C., Parazak, S. E., Budge, S., & Hilsenroth, M. (2018). Therapeutic alliance, empathy, and genuineness in individual adult psychotherapy: A meta-analytic review. Psychotherapy Research, 28(4), 593–605. 10.1080/10503307.2016.1204023

Nutt, D., & Carhart-Harris, R. (2021). The Current Status of Psychedelics in Psychiatry. JAMA Psychiatry, 78(2), 121. 10.1001/jamapsychiatry.2020.2171

NVivo Plus (Version 1.7.2). (2023). [Computer software]. Lumivero. www.lumivero.com

Passie, T., Loizaga-Velder, A., Danforth, A., Grob, C. S., Greer, G. R., Erritzoe, D., Oehen, P., Styk, J., Schlichting, M., Vermetten, E., Goksøyr, I. W., Palenicek, T., Mithoefer, M. C., Mithoefer, A., Anderson, B., Krediet, E., Gasser, P., Nielson, E. M., Gorman, I., … Guss, J. (2025). A model training curriculum for psychedelic, psycholytic, and entactogen-assisted psychotherapy. Journal of Psychopharmacology, 02698811241282759. 10.1177/02698811241282759

Phelps, J. (2017). Developing Guidelines and Competencies for the Training of Psychedelic Therapists. Journal of Humanistic Psychology, 57(5), 450–487. 10.1177/0022167817711304

Prusiński, T. (2022). The strength of alliance in individual psychotherapy and patient’s wellbeing: The relationships of the therapeutic alliance to psychological wellbeing, satisfaction with life, and flourishing in adult patients attending individual psychotherapy. Frontiers in Psychiatry, 13. 10.3389/fpsyt.2022.827321

R Core Team (2020). R: A language and environment for statistical computing. R Foundation for Statistical Computing, Vienna, Austria. https://www.R-project.org/.

Raison, C. L., Sanacora, G., Woolley, J., Heinzerling, K., Dunlop, B. W., Brown, R. T., Kakar, R., Hassman, M., Trivedi, R. P., Robison, R., Gukasyan, N., Nayak, S. M., Hu, X., O’Donnell, K. C., Kelmendi, B., Sloshower, J., Penn, A. D., Bradley, E., Kelly, D. F., … Griffiths, R. R. (2023). Single-Dose Psilocybin Treatment for Major Depressive Disorder: A Randomized Clinical Trial. JAMA, 330(9), 843. 10.1001/jama.2023.14530

Rosenbaum, D., Hare, C., Hapke, E., Herman, Y., Abbey, S. E., Sisti, D., & Buchman, D. Z. (2024). Experiential Training in Psychedelic-Assisted Therapy: A Risk-Benefit Analysis. Hastings Center Report, 54(4), 32–46. 10.1002/hast.1602

Sanders, J. W., & Zijlmans, J. (2021). Moving Past Mysticism in Psychedelic Science. ACS Pharmacology & Translational Science, 4(3), 1253–1255. 10.1021/acsptsci.1c00097

Sessa, B. (2012). The psychedelic renaissance: Reassessing the role of psychedelic drugs in 21st century psychiatry and society (pp. xi, 237). Muswell Hill Press.

Wampold, B. E. (2015). How important are the common factors in psychotherapy? An update. World Psychiatry, 14(3), 270–277. 10.1002/wps.20238

Watson, J., Steckley, P., & Evelyn, M. (2013). The role of empathy in promoting change. Psychotherapy Research : Journal of the Society for Psychotherapy Research, 24(3), 286–298. 10.1080/10503307.2013.802823

Yeh, S.-C. J., Chen, S.-H. S., Yuan, K.-S., Chou, W., & Wan, T. T. H. (2020). Emotional Labor in Health Care: The Moderating Roles of Personality and the Mediating Role of Sleep on Job Performance and Satisfaction. Frontiers in Psychology, 11. 10.3389/fpsyg.2020.574898

